# A Delphi study to establish a consensus definition and clinical reporting guidelines for Mesenchymal Stromal Cells

**DOI:** 10.1101/2021.06.18.21259151

**Authors:** Laurent Renesme, Kelly D Cobey, Maxime Lê, Manoj M Lalu, Bernard Thébaud

## Abstract

**Introduction:** Despite being more than two decades of research, Mesenchymal Stromal Cell (MSC) treatments are still struggling to cross the translational gap. Two key issues that likely contribute to these failures are i) the lack of clear definition for MSC and ii) poor quality of reporting in MSC clinical studies. To address these issues, we propose a modified Delphi study to establish a consensus definition for MSC and clinical reporting guidelines for MSC.

**Methods and analysis:** We will conduct a three-round international modified Delphi Survey. Findings from a recent scoping review examining how MSC are defined and reported in preclinical and clinical studies were used to draft the initial survey for round one of our Delphi. Participants will include a ‘core group’ of individuals as well as researchers whose work was captured in our scoping review. The core group will include stakeholders from different research fields including developmental biology, translational science, research methods, regulatory practices, scholarly journal editing, and industry. The first two survey rounds will be online, and the final round will take place in person. Each participant will be asked to rate their agreement on potential MSC definition characteristics and reporting items using a Likert scale. After each round, we will analyse data to determine which items have reached consensus for inclusion/exclusion, and then develop a revised questionnaire for any new items, or items that did not reach consensus.

**Ethics and dissemination:** This study received ethical approval from the Ottawa Health Research Network Research Ethics Board. To support the dissemination of our findings, we will use an evidence-based ‘integrated knowledge translation’ approach to engage knowledge users from the inception of the research. This will allow us to develop a tailored end-of-project knowledge translation plan to support and ensure dissemination and implementation of the Delphi results.

**Strengths and limitations of this study:**

- We proposed to address the current limitations in MSC experimental and clinical research with a rigorous and methodological consensus building method (Delphi method) that will allow for structured communication on controversial issues.
- To support dissemination and implementation of our results, we will engage stakeholders and end-users from the inception of the project – such as patient partners – and will develop a tailored end of project knowledge translation plan (integrated knowledge translation approach) in order to overcome historical issues related to community uptake.
- To address the main limitations of a Delphi method (e.g., lack of participation, no in-person interaction or information exchange), we use a modified Delphi survey with a Core group of stakeholders and a face-to-face meeting.

## INTRODUCTION

Since their original description^1^ and their first use as a therapeutic agent in humans,^2^ interest for Mesenchymal Stromal Cells (MSC) from the scientific and patient community keeps growing exponentially: a PubMed search with the query mesenchymal stromal cells between 1995 and 2021 found 73,876 results; more than half of the results were from the last 5 years. From a clinical research perspective, over 1300 MSC clinical trials have been registered on clinicaltrials.gov.^3^ Despite promising results of MSC in different preclinical diseases models, clinical trials using MSC in various medical conditions failed to deliver encouraging results.^4,5^ Many potential explanations for this disparity between preclinical and clinical results exist, including MSC characteristics, cell manufacturing processes, administration protocols or study participants’ characteristics such as disease severity and associated comorbidities.^6^ In addition, MSC characteristics (e.g., definition, characterization, immune compatibility, cell viability and dose) raise two main issues of i) lack of consensus definition for MSC and ii) broad variability in MSC characteristics reported. For example, a report from the Food and Drug Administration showed a significant heterogeneity in MSC products used in the clinical trials, with important differences in cell surface marker characterisation, product bioactivity assessment, as well as tissue sourcing and product manufacturing^7^. Regarding reporting of MSC characteristics, members of our group have performed systematic reviews of published MSC clinical trials and have found extremely poor (i.e., incomplete) reporting of cell products used.^8–10^ These observations have pushed some scientists to call to “clear up this stem cell mess”.^4^

The first important initiative to develop a consensus definition for MSC was provided by the International Society for Cell & Gene Therapy (ISCT), which in 2006 established the minimal criteria for human bone marrow-derived MSC and these were updated in 2019.^11,12^ These criteria were determined through a group decision-making method (e.g., informal consensus of a small number of experts), which is a consensus development method with several limitations.^13^ However, implementation of this MSC definition has been inconsistent and several researchers have expressed scepticism and highlighted limitations (e.g., cell surface markers phenotype, misinterpretation of differentiation assays, limitations in functionally defining stromal cells, etc.).^4,11,14^ For example, a scoping review conducted by our group to describe how MSC are defined and characterized in both preclinical and clinical research showed that only 18% of the articles from our sample explicitly referred to the ISCT criteria, and only 7% of clinical studies selection reported the 3 minimal criteria from the 2006 definition. The uncertainty and lack of consensus on this crucial issue of how to define MSC has also allowed questionable for-profit business to sell unproven and unlicensed “stem cell” therapies to patients and public.^15,16^

Therefore, there is an urgent need to develop a consensus definition of MSC and reporting guidelines to create standards for complete and transparent reporting of both cell characteristics and clinical trial/manufacturing details. A clear definition and reporting guidelines will be the cornerstones of a more robust, reproducible and transparent research, and are mandatory to better understand underlying factors that may contribute to efficacy (or a lack of efficacy) of MSC. Moreover, for patients and members of the public, having a clear definition of MSC will allow them to make informed decisions into which related treatments, clinical trials, or products they elect to partake in.

To address limitations of previous attempts to define MSC and support the dissemination and implementation of MSC definition and reporting guidelines, our research protocol is based on a modified Delphi method combined with an integrated knowledge translation approach.

The objectives of the current study are to: 1) develop, disseminate and implement an updated consensus definition of minimal criteria to define MSC; and 2) develop reporting guidelines for the clinical trials of MSC therapy. The work is not hypothesis testing and therefore, we have no a priori predictions related to study outcomes.

## METHODS AND ANALYSIS

### Ethics and dissemination

#### a. Ethics

Ethics approval was obtained from the Ottawa Health Research Network Research Ethics Board (REB Protocol ID# 20210187-01K).

#### b. Integrated knowledge translation (iKT) approach

The iKT will ensure that the MSC research community is aware of the recommendations described in our consensus definition and reporting guidelines in order to maximize the impact of the work. This entails the early inclusion of key stakeholders who have the ability to implement our recommendations, including patient partners. This participation begins from project inception and continues through to dissemination and implementation of study findings. The goal of the iKT approach is to ensure that the needs and preferences of stakeholders are considered throughout the project, with the idea being that successful partnership upstream will facilitate improved implementation and uptake downstream.^17^

Finally, to further support the dissemination of our findings (definition and reporting guidelines), we will use a structured approach to end-of-project, modeled on CIHR’s Knowledge Translation planning guide (https://cihr-irsc.gc.ca/e/45321.html). This essentially will involve considering the specific project findings and determining the dissemination goal (e.g., to increase awareness of the findings, to increase knowledge, to influence practice or policy), identifying key audiences, crafting messages tailored to specific audiences, and using strategies and media to reach each audience. A written KT plan will help ensure study outputs are effectively used. Our knowledge users will help interpret the Delphi results and craft dissemination messages. We plan to create an extension to existing clinical study reporting guidelines (e.g., CONSORT) that will focus on aspects unique to MSC therapy trials (e.g., manufacturing, characterization, storage and delivery of cells). To support its implementation, the guideline draft will be published as a preprint to obtain feedback from the scientific community, and associated explanatory documents such as a guidance development statement and an Explanation and Elaboration document will be provided.

#### c. Registration and Data Availability

This study protocol was registered using the Open Science Framework (OSF) (https://osf.io/3dsqx/). Data and study materials will be made publicly available at the time of publication using OSF.

#### d. Patient and public involvement

A Patient Partner is involved in our project since inception. The Patient Partner will provide feedback on the consensus definition of MSC and reporting guidelines development and on knowledge translation strategies. They will help to co-develop a communication plan for the general public by supporting the development of non-technical summaries for the scoping review and the Delphi findings as well as their dissemination to general public via patients’ associations and social media.

### Project overview

We will conduct a three-round Delphi survey. The Delphi method is a process that uses several rounds of surveying in order to reach consensus on a topic.^13,18^ Between rounds, responses to survey items are aggregated – items that reach a priori threshold for inclusion/exclusion are omitted, and any remaining or new items are shared with participants in the subsequent round to reconsider and vote on again. The Delphi method addresses group decision-making method’s limitations such as (i) group influences on individual performance, (ii) adequately accounting for divergent opinions, (iii) limiting the number of participants, (iv) meeting organisation and costs, (v) time limits for very complicated decisions, (vii) peer pressure and influence in the decision-making process, and (viii) logistical issues.^13^ The Delphi method has been successful in the past in solving contentious issues.^19–21^

### Participants

Participants for this Delphi study will be identified in two ways:

a. First, we will invite members of a ‘core group’ of experts that have been identified for the purpose of planning and implementing this research program. The core group members are diverse in their makeup, including in vitro, preclinical, and clinical researchers’ and include member with backgrounds in developmental biology, translational science, research methods, regulatory practices, scholarly journal editing, and industry.
b. Secondly, we will invite a ‘researcher panel’ to participate in the Delphi. This panel will be comprised of 311 researchers that we identified in our previous scoping review of MSC research conducted between March and May 2020 (submitted). Our scoping review (https://osf.io/3dsqx/) was developed to describe how MSC are currently defined in preclinical and clinical research, which will provide valuable themes to consider as items for the current Delphi survey. We extracted the corresponding author’s name, e-mail, and country of primary affiliation, from all original articles included in this scoping review. These authors will be contacted by a member of the research team and invited to participate in the Delphi survey.

### Recruitment

Participants on the Core group and on the Researcher panel will be approached for participation in the study using a standardized e-mail recruitment script (see Appendix 1). This script will provide participants with more information about the study and provide a link to access the informed consent form and the round one online Delphi survey. The Core Group will take part in all three rounds of the Delphi. The Researcher panel will be invited to the initial round only. This approach was taken to maximize the initial reach of our survey while maintaining a manageable number of individuals to facilitate our round three in person consensus meeting.

Participants will be given 3 weeks to complete each round of the Delphi survey. Reminder emails will be sent 7 and 14 days following the dissemination of each online questionnaire.

### Study Design

Our modified Delphi study will involve three rounds of surveying; the first two will take place via an online survey, while the third round will take place via an in-person consensus meeting. The study design is presented in Figure 1. The online Delphi surveys will be administered using Surveylet® (https://calibrum.com) a cloud-based platform specifically designed for Delphi surveys.

**Figure 1.**
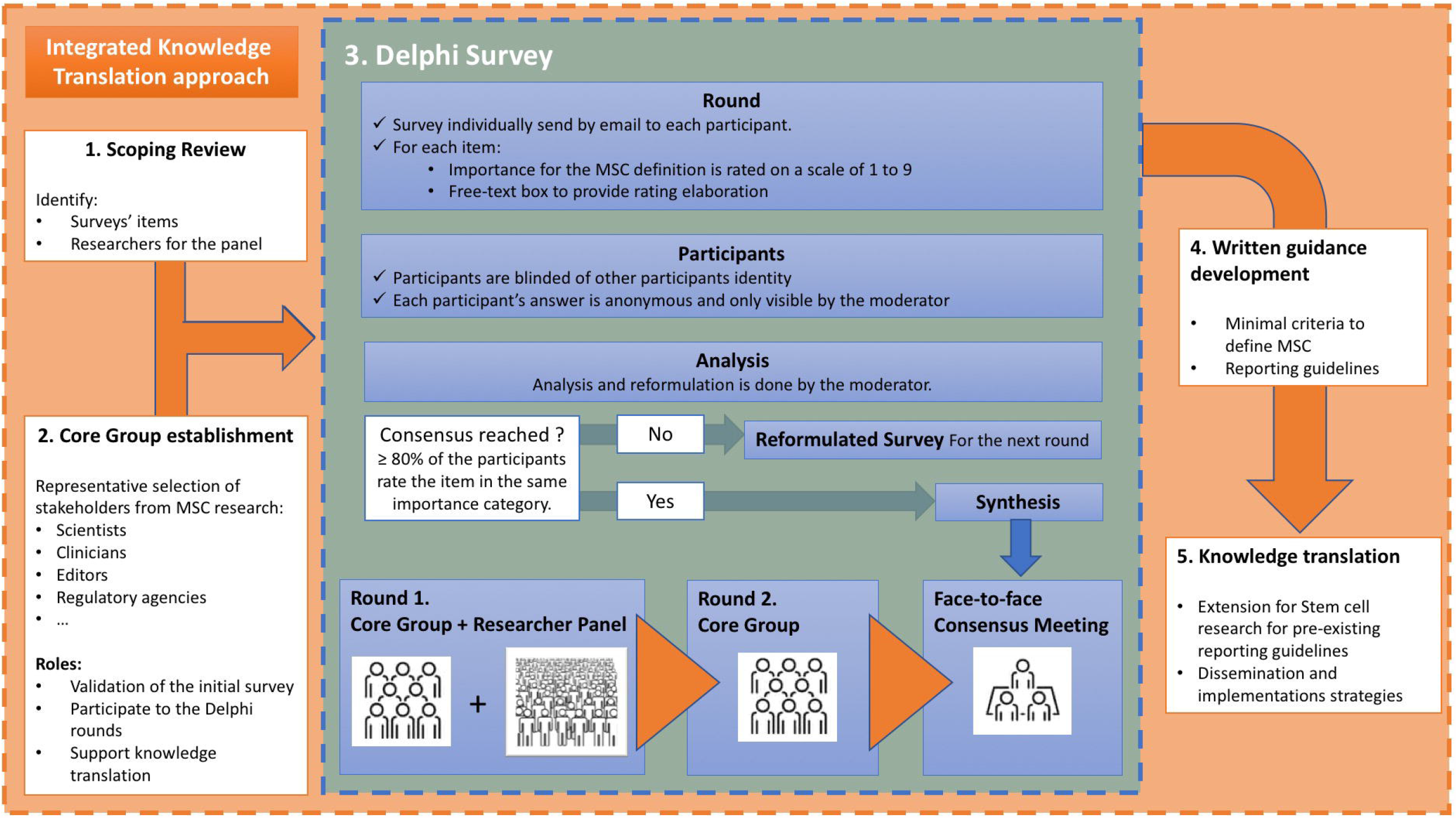
Study design. MSC: Mesenchymal stromal cell.

- Round 1 (online): completed by the Researcher panel and the Core group.
- Round 2 (online): completed by the Core group only.
- Round 3 (virtual consensus meeting): All items will be discussed by the Core Group members during a virtual satellite meeting at the 2021 TMM Meeting during a dedicated half-day.

### Delphi survey

We have generated an initial survey (https://osf.io/u579g/) which has three sections: i) participant demographics, ii) items for MSC definition, iii) items for reporting guidelines. Based on participants’ self-identified field of expertise, they will access different parts of the survey: MSC definition, reporting guidelines or both. This survey was piloted initially by six members of our research group. Then, the questionnaire was sent to eight leaders of opinion in MSC research (core group members and leadership within international stem cell scientific societies) to ensure relevance and completeness of our items for definition and reporting guidelines.

After completing the demographics section, participants will be asked to rate each survey item’s importance for MSC definition and/or as a minimal reporting criterion (e.g., ‘*A description of MSC capacity to adhere to a plastic surface when maintained in standard culture condition, is essential to define them*.‘) on a scale of 1 (strongly disagree) to 9 (strongly agree). A free-text response option will accompany each listed item to allow participants to elaborate on their rating. At the end of the survey, participants will also have the opportunity to suggest any new items that were not already listed on the survey. After each round, data from the survey will be aggregated. A revised survey will be developed for the subsequent that incorporates any newly suggested items, and items that did not reach consensus. Items that did not reach consensus will be presented with verbatim feedback participants made in the previous rounds.

### Data Analysis

#### a. Data analysis

Data will be analyzed using Excel. Data will be collected anonymously. The total number of participants, item rating scores, and demographic data will be summarized using frequencies and percentages. For each Likert scale item, we will report mean and range. Mean rating scores for each item will be categorized into three groups (unessential, potentially essential, essential). A median rating of 1-3 means an item is deemed unessential, 4-6 implies that an item is potentially essential, and a median rating of 7-9 is deemed essential to report.

Consensus will be achieved when at least 80% of Delphi participants rate the item in the same category of importance: unessential or essential. This threshold was described as the most commonly used in a systematic review of consensus in Delphis.^22^ Items that achieve consensus, as being ‘essential or unessential’ will be labelled as such, and in subsequent rounds participants will have the opportunity to comment on these items. Items that did not achieve consensus, achieve consensus as ‘potentially essential’, had major wording changes, or were added by Delphi participants will automatically move on to the subsequent rounds. For each round, response rate will be recorded and reported.

#### b. First round analysis

Any items from round 1 with disagreement between core group and researcher panel will be provided to the core group in round 2 to re-vote on. For transparency, all the results will be reported separately in a final manuscript so that any discrepancies between the core group and the research group will be clearly stated.

#### c. Second round analysis

After the second round with the core group, items that reach consensus (>80% agreement) in the essential category will be considered for inclusion in the MSC definition and the reporting guidelines. Items that reach consensus in the unessential category will be considered for exclusion. All remaining items that have not yet reached consensus will be presented and discussed at the final face-to-face virtual meeting where they will then be electronically voted on anonymously using Surveylet® software (https://calibrum.com).

### Potential limitations and mitigation strategies

One feature of the traditional Delphi method is that the participants are isolated from each other, with no in-person interaction or information exchange to limit group influences and peer pressure. Face-to-face interaction can be useful to identify reasons for any disagreements. To address this limitation, we plan to organize the third round as face-to-face virtual meeting, after which participants again vote anonymously. A lack of participation overall is another concern for any survey-based study. We have mitigated this risk by involving thought leaders from MSC science and other stem cell areas who have committed to contributing. Furthermore, our previously conducted scoping review captured 311 MSC researchers, and as such if not all researchers respond we still anticipate sufficient recruitment numbers. Another potential limitation of the Delphi method is respondent attrition between rounds. We have addressed this by engaging our core group from project inception. During the rounds, reminder emails will be sent to solicit greater responses, and to limit participant attrition between rounds, we plan to analyze the results of each round and provide feedback and a new questionnaire rapidly to maintain the interest and engagement of participants. Respondent attrition per round will be recorded and reported.

## CONCLUSION

There is a need to provide a clear definition of MSC and to create reporting guidelines to support more transparent, reproducible and ethical MSC research. Previous attempts to deliver a universally accepted definition of MSC have not been widely accepted or implemented by the community, mostly due to the difficulty of bringing stakeholders with divergent points of view to the same table and in implementing final products successfully. In collaboration with our core group of experts, we propose a methodologically sound approach for consensus development to address this gap. This project brings methods specific to meta-science and applies them within the MSC research community.

## Author contributions

LR: Conceptualization, methodology, and writing-original draft preparation.

MML, KC, BT: Conceptualization, funding acquisition, methodology, supervision and writing-review & editing.

ML (Patient Partner): Manuscript review and editing.

## Acknowledgements

The authors want to thanks the following members from the Thebaud lab for their valuable feedbacks on the initial Delphi questionnaire: Pauline Bardin, Chanele Cyr-Depauw, Flore Lesage, Marissa Lithopoulos, Ivana Mizikova and Liqun Xu.

## Appendix 1. Recruitment emails

### A. Core group

#### Recruitment Email

##### Subject Line: Invitation to contribute to MSC community consensus

Dear colleague,

You are being asked to participate in a Stem Cell Network funded research study that we are conducting at the Ottawa Hospital Research Institute, and that is led by Drs. Bernard Thébaud, Kelly Cobey and Manoj Lalu. The study name is: *Clearing-up the stem-cell-mess: A Delphistudy to establish and consensus: definition and reporting guidelines for MSC*.

The aim of our study is to reach consensus opinion on items that should be used to define Mesenchymal stromal or stem cells (MSC) for *in vitro*, preclinical *in vivo* animal experiments and clinical research. As such, we are conducting a Delphi survey to generate an international consensus on these issues. A scoping review of research on MSC was done to inform the current study items.

We are recruiting a Core group of experts composed by representative stakeholders from different fields of MSC research (developmental biology, translational scientists, clinical trialists, methodologists, journal editors, regulatory agencies representatives…). We have identified you as a leader of opinion in your field and are inviting you to join our Core group and participate in this study. We would value your expert opinion.

Briefly, your participation will consist of the completion of two online surveys on SurveyLet (calibrum.com) and a virtual face-to-face meeting. For the online surveys, you will be asked to rate items for consideration based on level of importance on a scale from 1 to 9. You will also be provided with the opportunity to submit a free-text response for each item. The survey will be open for 3 weeks, after which the link will expire. Reminders emails will be sent 7 and 14 days following the administration of the survey. The 2 online surveys will be administered with a 3-weeks interval in between. The virtual face-to-face meeting will be held as a satellite conference at the 2021 Till & McCulloch meetings in November 2021

##### Conflicts of Interest

There are no conflicts of interest to declare.

##### Voluntary Participation and Withdrawal

Participation is voluntary. You can withdraw from participating at any time while completing the survey by closing your browser; however, once the completed survey has been submitted, it will not be possible to withdraw your information. Any information recorded before you withdraw will be used by the researchers for the purposes of the study, but no information will be collected after you withdraw your permission.

##### Risks and/or Benefits

Participation involves no risks to you. You may not receive direct benefit from participating in this study. We hope the information learned from this study will help to develop, disseminate and implement consensus definition for MSC and reporting guidelines for MSC clinical studies.

##### Privacy/Confidentiality

Records identifying you at this center will be kept confidential and, to the extent permitted by the applicable laws, will not be disclosed or made publicly available, except as described in this consent document.

Authorized representatives of the following organizations may look at your original (identifiable) research records at the site where these records are held, to check that the information collected for the study is correct and follows proper laws and guidelines.

- The Ottawa Hospital Research Institute, the Sponsor of this study
- The Ottawa Health Science Network Research Ethics Board
- The Children’s Hospital of Eastern Ontario Research Ethics Board
- The Children’s Hospital of Eastern Ontario – Ottawa Children’s Treatment Centre and the Research Institute

Information that is collected about you for the study (called study data) may also be sent to the organizations listed above.

This study requires the transfer of identifiable information to Calibrum, Inc, whose servers are located in the United States, for the purposes of on-line survey administration and data analysis. The following information will be transferred: Email Address.

Communication via e-mail is not absolutely secure. We do not recommend that you communicate sensitive personal information via e-mail.

If the results of this study are published, your identity will remain confidential. It is expected that the information collected during this study will be used in analyses and will be published and presented to the scientific community at meetings and in journals.

Even though the risk of identifying you from the study data is very small, it can never be completely eliminated.

Any information sent outside of Canadian borders may increase the risk of disclosure of information because the laws in those countries dealing with protection of information may not be as strict as in Canada. The information will be transferred in compliance with all relevant Canadian privacy laws. By returning your completed survey, you are consenting to the disclosure of your information to organizations located outside of Canada.

##### Cost and/or Payment

You will not be paid for being in this study, nor will there be any cost to you.

##### Rights of Participants

You have the right to be informed of the results of this study once the entire study is complete. If you would like to be informed of the results of this study, please let the research team know.

Your rights to privacy are legally protected by federal and provincial laws that require safeguards to ensure that your privacy is respected.

##### Questions

If you have any questions about taking part in this study, please contact me at, or the Principal Investigator Dr. Bernard Thébaud at.

If you have questions about your rights as a participant or about ethical issues related to this study, you can talk to someone who is not involved in the study at all. Please contact The Ottawa Health Science Network Research Ethics Board, Chairperson at 613-798-5555 extension 16719.

##### Implied Consent

By completing the survey, your consent to participate is implied.

If you would like to participate in this study, please click on the link below to access the survey: **[insert link]**.

Thank you,

Laurent Renesme, MD

The Ottawa Hospital Research Institute

### B. Researcher panel

#### Recruitment Email

##### Subject Line: Invitation to contribute to MSC community consensus

Dear colleague,

You are being asked to participate in a Stem Cell Network funded research study that we are conducting at the Ottawa Hospital Research Institute, and that is led by Drs. Bernard Thébaud, Kelly Cobey and Manoj Lalu. The study name is: *Clearing-up the stem-cell-mess: A Delphistudy to establish and consensus: definition and reporting guidelines for MSC*.

The aim of our study is to reach consensus opinion on items that should be used to define Mesenchymal stromal or stem cells (MSC) for *in vitro*, preclinical *in vivo* animal experiments and clinical research. As such, we are conducting a Delphi survey to generate an international consensus on these issues. We conducted a scoping review of research on MSC to inform the current study, we identified you and are inviting you to participate in this study because we retrieved an article you published using MSC in our scoping review. We would value your expert opinion.

Briefly, your participation will consist of the completion of one online survey on SurveyLet (calibrum.com). You will be asked to rate items for consideration based on level of importance on a scale from 1 to 9. You will also be provided with the opportunity to submit a free-text response for each item. The survey will be open for 3 weeks, after which the link will expire. Reminders emails will be sent 7 and 14 days following the administration of the survey.

##### Conflicts of Interest

There are no conflicts of interest to declare.

##### Voluntary Participation and Withdrawal

Participation is voluntary. You can withdraw from participating at any time while completing the survey by closing your browser; however, once the completed survey has been submitted, it will not be possible to withdraw your information. Any information recorded before you withdraw will be used by the researchers for the purposes of the study, but no information will be collected after you withdraw your permission.

##### Risks and/or Benefits

Participation involves no risks to you. You may not receive direct benefit from participating in this study. We hope the information learned from this study will help to develop, disseminate and implement consensus definition for MSC and reporting guidelines for MSC clinical studies.

##### Privacy/Confidentiality

Records identifying you at this center will be kept confidential and, to the extent permitted by the applicable laws, will not be disclosed or made publicly available, except as described in this consent document.

Authorized representatives of the following organizations may look at your original (identifiable) research records at the site where these records are held, to check that the information collected for the study is correct and follows proper laws and guidelines.

- The Ottawa Hospital Research Institute, the Sponsor of this study
- The Ottawa Health Science Network Research Ethics Board
- The Children’s Hospital of Eastern Ontario Research Ethics Board
- The Children’s Hospital of Eastern Ontario – Ottawa Children’s Treatment Centre and the Research Institute

Information that is collected about you for the study (called study data) may also be sent to the organizations listed above.

This study requires the transfer of identifiable information to Calibrum, Inc, whose servers are located in the United States, for the purposes of on-line survey administration and data analysis. The following information will be transferred: Email Address.

Communication via e-mail is not absolutely secure. We do not recommend that you communicate sensitive personal information via e-mail.

If the results of this study are published, your identity will remain confidential. It is expected that the information collected during this study will be used in analyses and will be published and presented to the scientific community at meetings and in journals.

Even though the risk of identifying you from the study data is very small, it can never be completely eliminated.

Any information sent outside of Canadian borders may increase the risk of disclosure of information because the laws in those countries dealing with protection of information may not be as strict as in Canada. The information will be transferred in compliance with all relevant Canadian privacy laws. By returning your completed survey, you are consenting to the disclosure of your information to organizations located outside of Canada.

##### Cost and/or Payment

You will not be paid for being in this study, nor will there be any cost to you.

##### Rights of Participants

You have the right to be informed of the results of this study once the entire study is complete. If you would like to be informed of the results of this study, please let the research team know. Your rights to privacy are legally protected by federal and provincial laws that require safeguards to ensure that your privacy is respected.

##### Questions

If you have any questions about taking part in this study, please contact me at, or the Principal Investigator Dr. Bernard Thébaud at.

If you have questions about your rights as a participant or about ethical issues related to this study, you can talk to someone who is not involved in the study at all. Please contact The Ottawa Health Science Network Research Ethics Board, Chairperson at 613-798-5555 extension 16719.

##### Implied Consent

By completing the survey, your consent to participate is implied.

If you would like to participate in this study, please click on the link below to access the survey: **[insert link]**.

Thank you,

Laurent Renesme, MD

The Ottawa Hospital Research Institute

